# The rapid identification and diagnosis of meniscus tear by Magnetic Resonance Imaging using a deep learning model

**DOI:** 10.1101/2022.01.11.22269112

**Authors:** Jie Li, Kun Qian, Jinyong Liu, Zhijun Huang, Yuchen Zhang, Guoqian Zhao, Huifen Wang, Meng Li, Xiaohan Liang, Fang Zhou, Xiuying Yu, Lan Li, Xingsong Wang, Xianfeng Yang, Qing Jiang

**Author notes:** Corresponding to Prof. Xingsong Wang, School of Mechanical Engineering, Southeast University, No.2 Southeast University Road, Nanjing, China, 210000. E-mail address Prof. Xianfeng Yang, Department of Radiology, Drum Tower Hospital affiliated to Medical School of Nanjing University, No.321 Zhongshan Road, Nanjing, China, 210000. These authors contributed equally to the work (Jie Li, Kun Qian, Jinyong Liu).

## Abstract

**Objective:** The meniscus tear is a common problem in sports trauma. The imaging diagnosis mainly depends on the MRI. To improve the diagnostic accuracy and efficiency, a deep learning model was employed in this study and the identification efficiency has been evaluated.

**Methods:** The standard knee MRI images of 924 individual patients were used to complete the training, validation, and testing process. The Mask R-CNN was considered as the deep learning network structure, and the ResNet50 was considered as the backbone network. The deep learning model was trained and validated with a dataset containing 504 and 220 patients, respectively. The accuracy testing was performed on a dataset of 200 patients and reviewed by an experienced radiologist and a sports medicine physician.

**Results:** After training and validation, the deep learning model effectively recognized the healthy and injured meniscus. The overall average precision of the bounding box and pixel mask was more than 88% when the IoU threshold value was 0.75. The detailed average precision of three types of menisci (healthy, torn, and degenerated) was ranged from 68% to 80%. The overall sensitivity of the bounding box and pixel mask was more than 74% at the IoU threshold from 0.50 to 0.95. The diagnosis accuracy for the healthy, torn, and degenerated meniscus was 87.50%, 86.96%, and 84.78%, respectively.

**Conclusion:** The Mask R-CNN recognized effectively and predicted the meniscus injury, especially for the tears that occurred at different parts of the meniscus. The recognition accuracy was admirable. The diagnostic accuracy can be further improved with the increase of the training sample size. Therefore, this tool has great potential in the application for the diagnosis of meniscus injury.

**The translational potential of this article:** Deep learning model has unique effect in reducing doctors’ workload and improving diagnosis accuracy. It can identify and classify injured and healthy meniscus more accurately after training and learning datasets. The torn and degenerated meniscus can also be distinguished by this model. This technology could serve as an effective tool for clinical MRI-assisted diagnostics in meniscus injury.

## 1. Introduction

The meniscus is commonly referred to as a fibrocartilaginous structure located in the knee joint cavity, between the femur and tibia. According to the position of the meniscus, it can be divided into medial meniscus and lateral meniscus. They provide strength to the joint and absorb the impact forces to protect it [1, 2]. Meniscus injuries can occur from the destruction of the meniscus integrity by various conditions such as dysplasia, chronic strain, and acute sprain. These injuries lead to a series of clinical symptoms such as pain and dysfunction. Meniscus injuries are very common, with an incidence rate of 6 to 7 in 10000 [3], which seriously affects the mobility and quality of life of the patients. Once the diagnosis is confirmed, most of the cases need surgical treatment. Accurate and timely preoperative diagnosis is of great significance.

The MRI generates high soft-tissue imaging resolution. This method can clearly visualize the shape and internal structure of the meniscus. Therefore, it is the preferred method of examination for the diagnosis of meniscus injuries [6, 11]. The meniscus produces a uniform low signal on magnetic resonance imaging (MRI) sequences, and the FS FSE PDWI (fat-suppressed fast spin-echo proton density-weighted image) is the most commonly used in the meniscus injury diagnosis. A multi-center study on the meniscus injury showed great clinical significance to analyze the risk and prognosis of the meniscus injury [12]. However, the accuracy of the MRI diagnosis is limited, which is affected by several factors including 1) several different tissues are situated around the meniscus and the shape of these tissues is irregular, 2) the abnormal signal of a meniscus tear is very small and is not easy to be found on the image, 3) the amount of MRI data may be extremely huge in a multi-center study (each patient has about 100 images), 4) the doctors’ diagnostic level are different, and the accuracy of diagnosis is affected by the experience, and 5) other subjective factors.

In recent years, the application of artificial intelligence (AI) in the field of medical imaging has become a research hotspot, and researchers believe that AI has the potential to provide accurate diagnosis and treatment. Deep learning and other AI applications can effectively improve the efficiency of data processing and reduce human errors through repetitive learning to identify disease patterns [13, 14]. Traditional machine learning algorithms mainly include neural network, k-nearest neighbor, support vector machine, naive Bayes classifier, and random decision forest. These algorithms rely on artificial intelligence’s shallow features. In deep learning, there is no need to specify the features manually. The machine can learn by itself through the data set training, which provides an advantage and brings a breakthrough in image processing.

Great progress has been made in the in-depth analysis of the knee MRI images using the AI, but it is far less used than in other critical conditions such as a tumor, nerve damage, and pulmonary nodule. Compared to the bone and cartilage, the study of the meniscus is limited, and image segmentation and post-processing are not feasible. Among the studies regarding meniscus tear and AI, most of the studies analyzed only the sagittal plane, and a small number of studies analyzed the sagittal plane, coronal plane, and cross-section [17]. The area under the curve of these studies ranged from 0.847 to 0.910 [18], meaning that this technology should be improved in the diagnostic accuracy of the MRI.

The slice thickness is an important parameter in the meniscus MRI examination. In the previous studies, the scanning layer thickness ranged from 0.7 mm to 3.0 mm [7, 11], which made the data sources lacking homogeneity. Based on the clinical practice, this study aimed to utilize the most commonly used sequence and scanning layer thickness in the model training, which has a wider application range and is beneficial for multi-center research in the future. After obtaining the feature map of the meniscus MRI images through convolutional neural networks, Mask R-CNN will perform classification, regression, and pixel-level mask diagnosis on original meniscus images. To verify the recognition accuracy of the deep learning network, the results were evaluated by experienced doctors and arthroscopic surgery. We anticipate that this technology could serve as an effective tool for clinical MRI-assisted diagnostics in meniscus injury.

## 2. Methods

### 2.1. Process of the MRI scanning

The studies followed relevant guidelines, and had approval from the ethics committee of Drum Tower Hospital affiliated to the Medical School of Nanjing University. All patients underwent MRI using a 3.0 Tesla MR imaging system (United Imaging Co., Ltd., Shanghai, China) with a dedicated knee coil in supine position and feet first. The sagittal fat-suppressed proton density-weighted (PDW) MR images were acquired digitally from the picture archiving and communication system (PACS; Neusoft Medical Systems Co., Ltd., Shenyang, China) in the Joint Photographic Experts Group (JPEG) format. The parameters of the MR FS PDW sequence were 3 mm slice thickness, 0.3 mm gap, 1500 ms time of repetition, 40 ms time of echo, 16×16 cm^2^ field of view, and 1 number of signal average. The sagittal position lines were set perpendicular to the line of the posterior femoral condyle on transverse images, and perpendicular to the articular surface of the tibial plateau on coronal images.

### 2.2. The inclusion criteria

According to the clinical diagnostic guidelines, the meniscus was divided into the meniscus without tear and with a tear. The diagnostic criteria of the meniscus tear were abnormal high signal in the meniscus, and the high signal involved at least in one articular surface of the meniscus or reached the free edge of the meniscus [19]. The data sources of the study were from the same MRI equipment and scanned by technicians who had the same standardized training experience and used the scanning parameters all the time. In the image processing stage, the images without motion artifacts or any other magnetic artifacts, which affected the meniscus were included.

### 2.3. The image dataset and masking

The meniscus MRI image dataset was retrieved and produced by combining clinical testing. For the recognition of fine results, the size of the acquired meniscus MRI images was selected as 1188×1372 pixels. The MRI images of 924 individual patients (18 images per patients) were collected and labeled to make the common objects in context datasets, in which 504 individual patients were used as the training dataset, 220 individual patients were used as the verification dataset, and 200 individual patients were used as the internal testing dataset. In addition, images of 180 individual patients from 8 hospitals were considered as external testing dataset. In order to visually observe the health of the menisci, the position and shape of the cartilage tissues were extracted and displayed.

The meniscus MRI images were manually segmented into 10 categories: cartilage tissue (CA), anterior horn tear (AH_tear), posterior horn tear (PH_tear), meniscus body tear (MBT), anterior horn degeneration (AD), posterior horn degeneration (PD), meniscus body degeneration (MBD), anterior horn intact (AH_intact), posterior horn intact (PH_intact), and meniscus body health (MBH). During the marking and labeling process, the cartilage without the full display has been discarded, and the pixels that could not be distinguished from health or injury were also ignored. **Figure 1** demonstrates the visualization process of the meniscus datasets. Usually, the cartilage was displayed clearly, and the healthy and injured meniscus was marked based on the doctor’s diagnosis.

**Figure 1.**
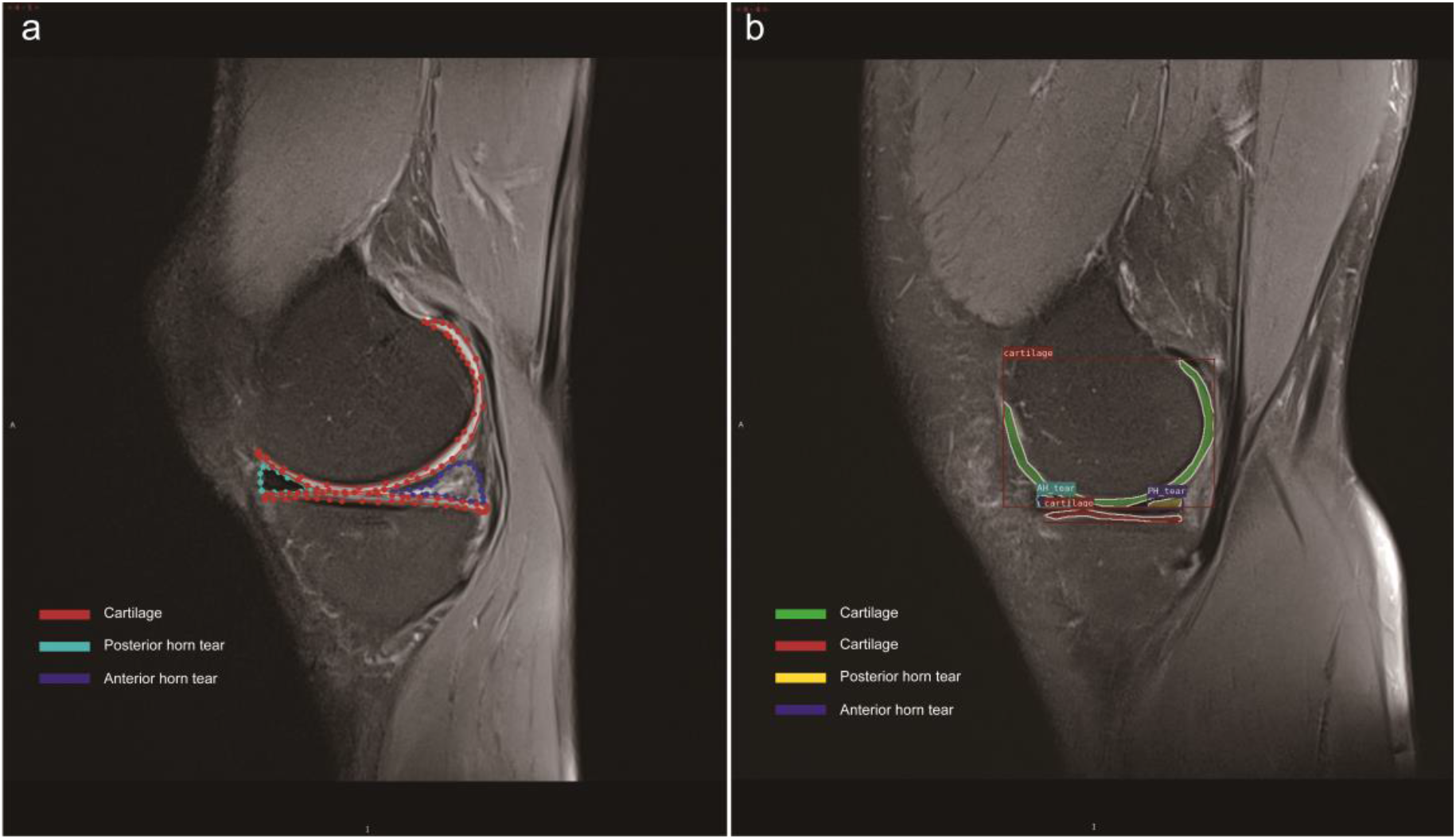
Meniscus MR Image dataset visualization process. (a) The marking process of the image, (b) The exported image from the deep learning model.

Due to the limitation of patient number, the number of images in the dataset used for training and verification may not be large enough. In the dataset establishment stage, data augmentation technology was used to supplement the collected dataset. Based on the labeled MRI image dataset, the geometric transformation, lighting adjustment, Gaussian filtering and noise addition were used to expand the number of samples in the data set. In order to inhibit the labeling errors, three geometric transformation methods were used: horizontal mirroring, vertical mirroring, and diagonal mirroring. By geometric transformation, one MRI image was transformed into four different images. The following processing was performed on these four images: Gaussian filtering, brightness enhancement, brightness reduction and adding noise (such as salt and pepper noise). The MRI images generated by the geometric transformation can simulate the difference caused by the slice angle and position. New images with different brightness can simulate different fat suppression level. Gaussian filtering blurred original MRI images and noise added more interference to images. As shown in **Figure 2**, through data augmentation and label processing, the dataset has been expanded by 20 times. The segmentation categories in the meniscus dataset are shown in **Table 1**. The total number of labels in the training dataset, validation dataset, and internal testing dataset was 30080, 16520, and 1012, respectively.

**Figure 2.**
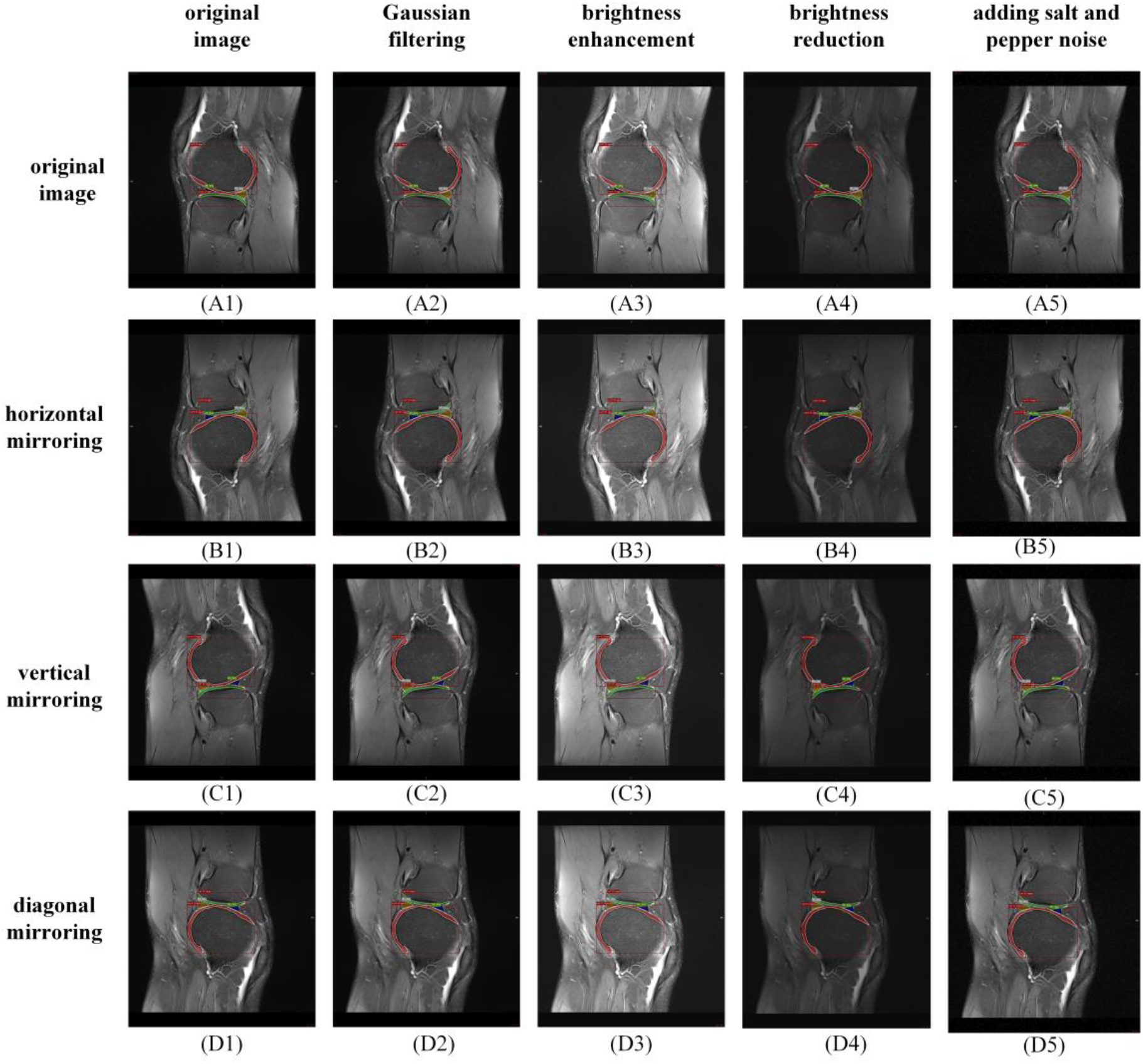
The illustration diagram of the dataset augmentation technique.

**Table 1.** Meniscus dataset and demographic breakdown.

|  | Patients number | CA | PT_tear | AT_tear | MB T | P D | A D | MB D | AH_intact | PH_intact | MB H | Total |
| --- | --- | --- | --- | --- | --- | --- | --- | --- | --- | --- | --- | --- |
| Training dataset | 504 | 19780 | 1620 | 860 | 560 | 780 | 820 | 380 | 2660 | 2080 | 540 | 30080 |
| Verification dataset | 220 | 7260 | 1260 | 840 | 300 | 420 | 700 | 240 | 3020 | 1980 | 500 | 16520 |
| Testing dataset | 200 | 348 | 114 | 65 | 33 | 50 | 56 | 22 | 164 | 129 | 31 | 1012 |

### 2.4. The network architecture

In this process, the Mask R-CNN was employed as the deep learning network structure to classify and segment the meniscus MRI images [20]. As shown in **Figure 3**, the process of deep learning for the identification of meniscus injuries mainly included two stages. The first stage was the generation of candidate regions, which primarily included the feature extraction by convolutional neural networks, Region Proposal Network (RPN) [21], and RoIAlign layer [20]. The second stage was the classification and regression of the objects and mask generation.

**Figure 3.**
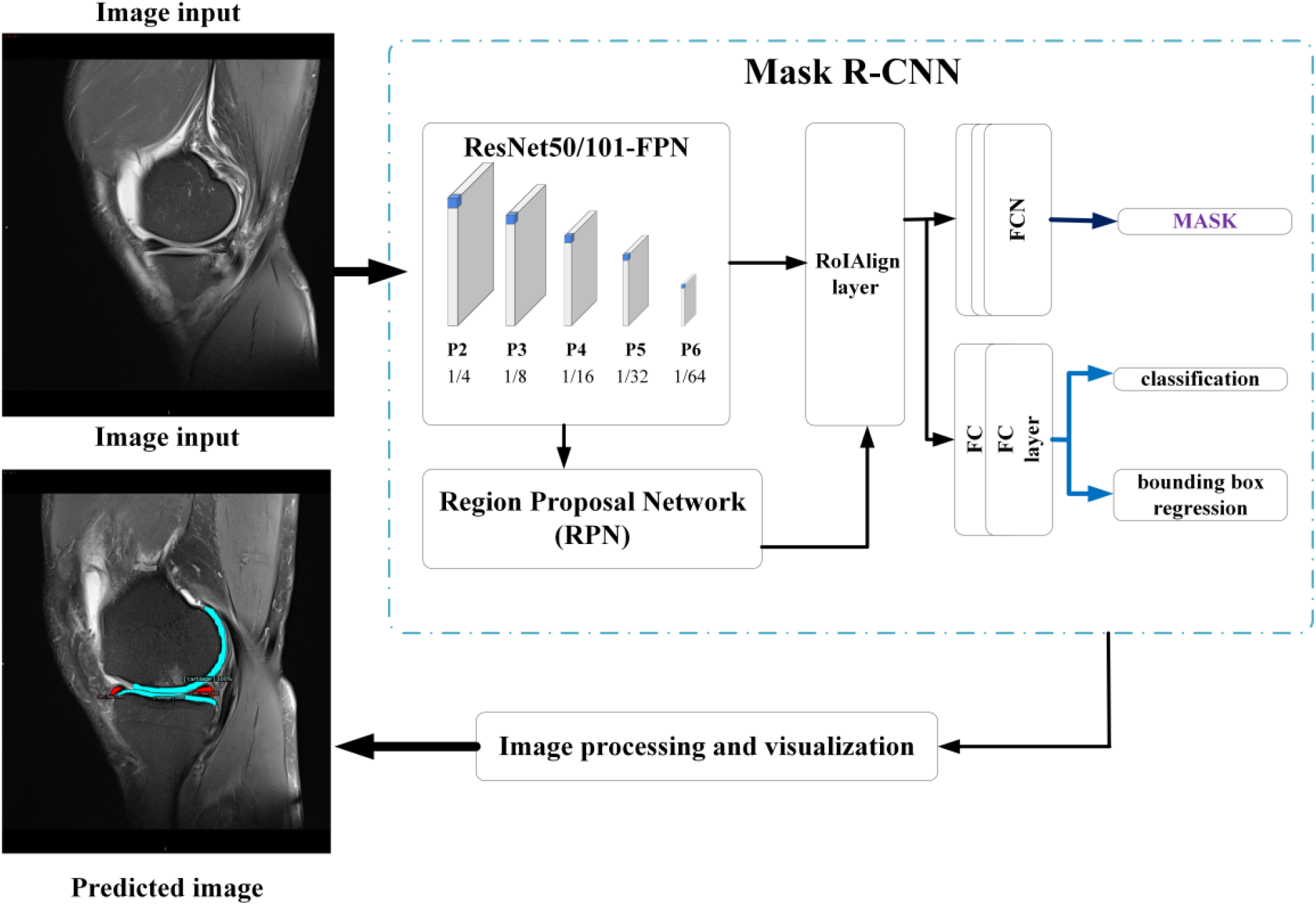
Architecture of the deep learning networks for the identification of torn menisci.

The feature map extraction was completed using the ResNet50 architecture as the backbone network. Because the ResNet50 had deeper network layers, it produced abundant feature information after the convolution and pooling of the original images. The ResNet50 combined with the Feature Pyramid Networks combined the feature maps from the bottom layer with the upper layer, which fully utilized the features of the different depths [22]. The purpose of the RPN was to recommend the region of interest (ROI) to the network. Briefly, the meniscus MRI image was inputted in the RPN, and the ROI of the original image was extracted by the 9-size anchor to output the region of recommendation score. The bilinear interpolation was used in the RoIAlign to extract the fixed-sized feature maps (for example, 7×7 pixel) from each ROI.

The Mask R-CNN finally outputted three branches of the meniscus images: classification, bounding box regression, and a mask branch (Fully Convolutional Networks) [23]. In the data set for the identification of meniscus injuries, the number of categories was 8 (background and other 7 categories), the output depth of the classification and regression network was 8 and the output mask network was 28×28×8.

### 2.5. Training

The ResNet50 has been adopted as the backbone network to train the meniscus MRI image datasets. The dataset was trained on a GPU (RTX 2070; NVIDIA, Santa Clara, CA, USA) for 10000 epochs, the initial learning rate was 0.01 (drop with training), the IMS_PER_BATCH was 2, and the NUM_CLASSES was 8. During the training process, the loss function was defined as:

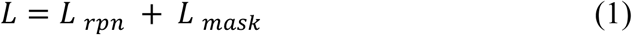

Where the *L* _*cls*_ and*L* _*box*_ respectively represented theclassification loss and bounding-box loss:

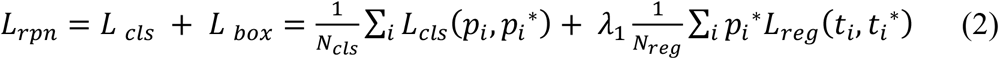

Where the *N* represented the number of corresponding anchors or bounding boxes; the hyper-parameters *λ* and *γ* balanced the training losses of the regression and mask branch. The *L* _*cls*_ represented the classification loss function and was expressed as:

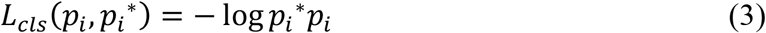

Where the *i* was the index of an anchor in a mini-batch; *p*_*i*_ was the predicted classification probability of anchor *i*; *p*_*i*_* represented the ground-truth label (correct and positive label) probability of the anchor *i*; *p*_*i*_* was 1 for the positive anchor and 0 for the negative anchor.

The *L* _*box*_ was bounding-box loss defined over a tuple of true bounding-box regression targets:

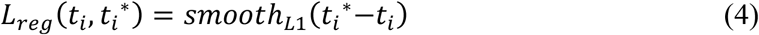

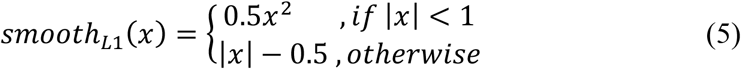

Where the *t*_*i*_* = (*t*_*x*_*, *t*_*y*_*, *t*_*w*_*, *t*_*w*_*) indicated the differences between the ground-truth label box and the positive anchor in four-parameter vectors (the horizontal and vertical coordinate values of the center point in the bounding box; the width and height of the bounding box); The *t*_*i*_ = (*t*_*x*_, *t*_*y*_, *t*_*w*_, *t*_*w*_) represented the difference between the diagnosis bounding box and the ground-truth label box:

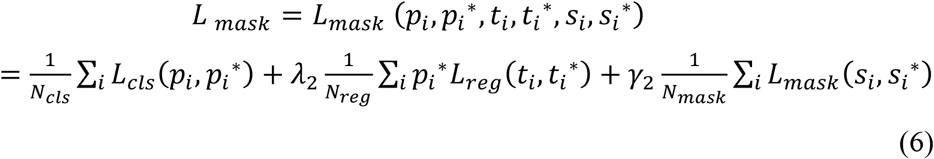

The definition of the *L* _*mask*_ allowed the network to generate masks for every class without the competition among classes. The *L* _*mask*_ was defined as the average binary cross-entropy loss used by a per-pixel sigmoid. The mask branch had a K*m*^2^ - dimensional output for each ROI (K was the number of classes). The *L* _*mask*_ was only defined on the k-th mask.

### 2.6. Model performances evaluation

To estimate the identification effect, the meniscus MRI image testing dataset was used for testing and evaluation. Intersection over Union (IoU) was used, which was the ratio of the intersection and union of the candidate bound area (C) and the ground truth bound area (G).

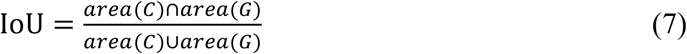

The formulas’ precision and recall were:

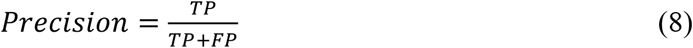

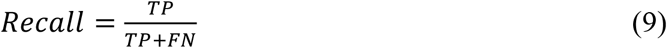

where the True Positive (TP) represented the resultant number of the IoU values greater than the threshold values (generally is 0.5). The False Positive (FP) represented the number of the IoU values less than the threshold values. The False Negative (FN) represented the number of unrecognized targets.

The Average Precision (AP) was used to measure the identification accuracy. For multi-class diagnosis, the AP was the average precision of multiple categories. The formula was:

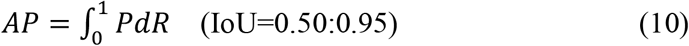

The AP50 and AP75 were APs when the IoU threshold was greater than 0.5 and greater than 0.75, respectively. The APs, APm, and APl were respectively represented as the AP for small objects (area <32^2^), medium objects (32^2^<area <96^2^), and large objects (96^2^<area).

### 2.7. Diagnosis accuracy evaluation

The accuracy of identification was evaluated by comparing the output results with the experienced radiologist. Briefly, 200 individual patients having meniscus tears, meniscus degeneration, and intact meniscus were identified as the internal testing dataset using the model. And 180 patients from 8 hospitals were regarded as external testing dataset. Among them, 90 patients (30 healthy patients, 30 patients with meniscus degeneration, and 30 patients with meniscus tear) were scanned using a 1.5T MRI, and 90 patients (30 healthy patients, 30 patients with meniscus degeneration, and 30 patients with meniscus tear) were scanned using the 3.0T MRI, the scanning parameters were listed in **Table 2**. The output results were assessed by an experienced radiologist and sports medicine physician. Additionally, 40 patients with meniscus tear which diagnosed by arthroscopic surgery were randomly selected to verify the diagnostic accuracy of this model.

**Table 2.** MR imaging system and scanning parameters.

| Types | Model | Field strength | MR sequence | Field of view | Time of repetition | Time of echo | Slice Thickness | Matrix |
| --- | --- | --- | --- | --- | --- | --- | --- | --- |
| Philips | Intera | 1.5 | FS-T2W | 18cm*18cm | 1800ms | 30ms | 4mm | 200*160 |
| United Imaging | uMR790 | 3.0 | FS-PDW | 16cm*16cm | 1500ms | 40ms | 3mm | 320*288 |
| Siemens | Skyra | 3.0 | FS-PDW | 17cm*19.6cm | 2600ms | 36ms | 3.5mm | 384*384 |
| Siemens | Avanto | 1.5 | FS-PDW | 16cm*16cm | 3000ms | 31ms | 4mm | 640*640 |
| Philips | Multiva | 1.5 | FS-PDW | 16cm*16cm | 2000ms | 25ms | 4mm | 288*224 |
| GE | Architect | 3.0 | FS-PDW | 16cm*16cm | 2500ms | 38ms | 4mm | 512*512 |
| Siemens | Avanto | 1.5 | FS-PDW | 22.2cm*16.6cm | 2000ms | 19ms | 4.5mm | 640*640 |
| Siemens | Skyra | 3.0 | FS-PDW | 16cm*16cm | 2800ms | 32ms | 3.5mm | 352*288 |
| GE | 750 | 3.0 | FS-PDW | 18cm*18cm | 1941ms | 35ms | 3.5mm | 352*224 |

## 3. Results

### 3.1. The Mask R-CNN training

The loss function and accuracy in the training process of the Mask R-CNN are shown in **Figure 4**. After 10000 iterations, the loss function was relatively low, and the accuracy increased to 0.96. More training and larger datasets were conducive to increasing accuracy and avoiding overfitting.

**Figure 4.**
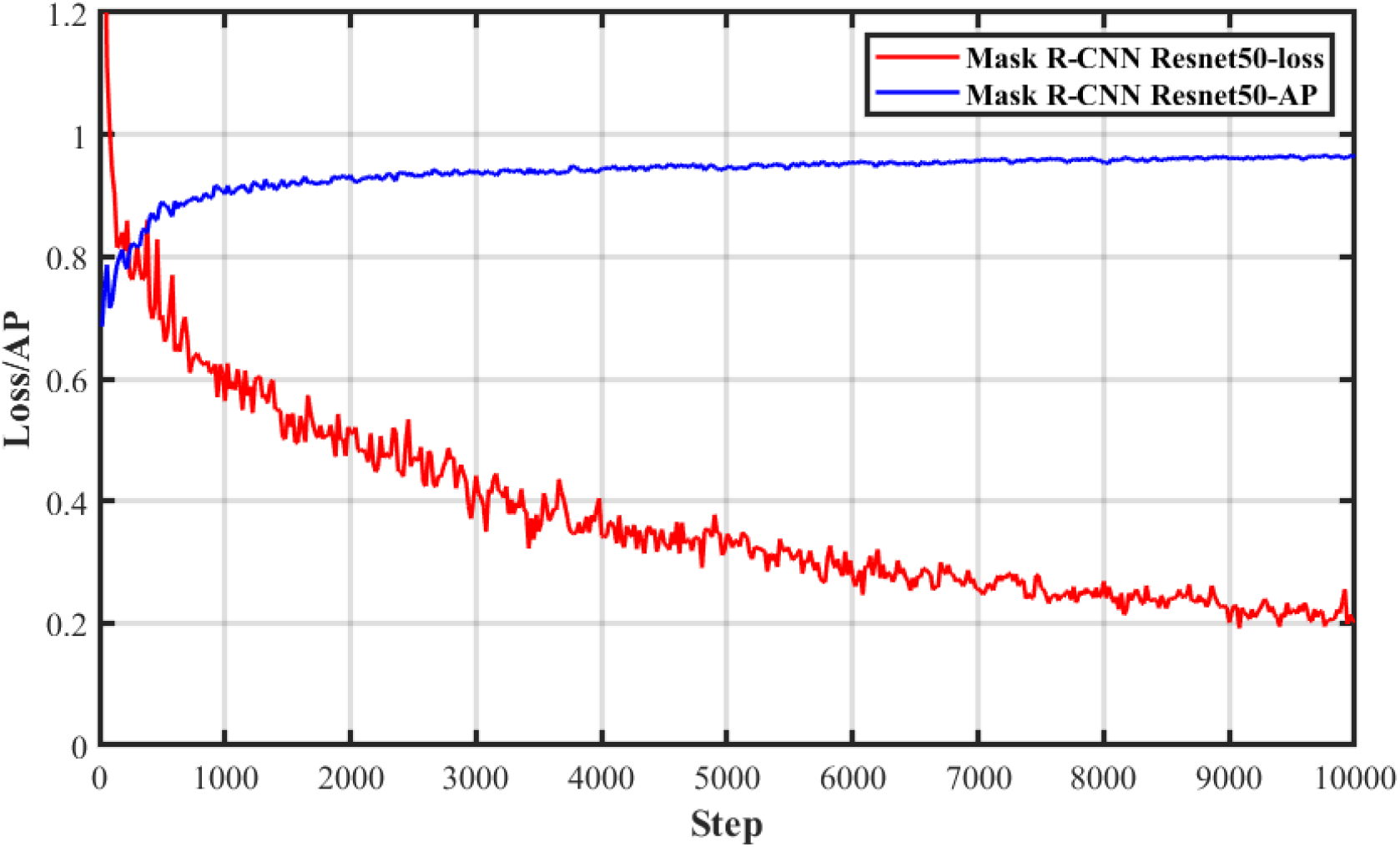
Loss function and accuracy in the training process of the Mask R-CNN.

### 3.2. The Image identification of meniscus

**Figure 5** shows the classification and instance segmentation of a meniscus MR image, and the targeted objects were marked by both bounding box and pixel. The reorganization diagnosis results of the Bbox and Mask are shown in **Figure 6** and **Figure 7**. The Bbox represented the bounding box containing the target objects (**Figure 6**). The Mask represented the predicted pixels of the tissues, like cartilage and meniscus (**Figure 7**). Different colors were employed to distinguish the cartilage and meniscus: Watchet blue bounding boxes and pixels represented the cartilage tissue (CA), red bounding boxes and pixels represented the injured meniscus (PT, AT, and MBT), yellow bounding boxes and pixels represented the degenerated meniscus (AD, MBD, and PD), and green bounding boxes and pixels represented the healthy meniscus (AH, MBH, and PH).

**Figure 5.**
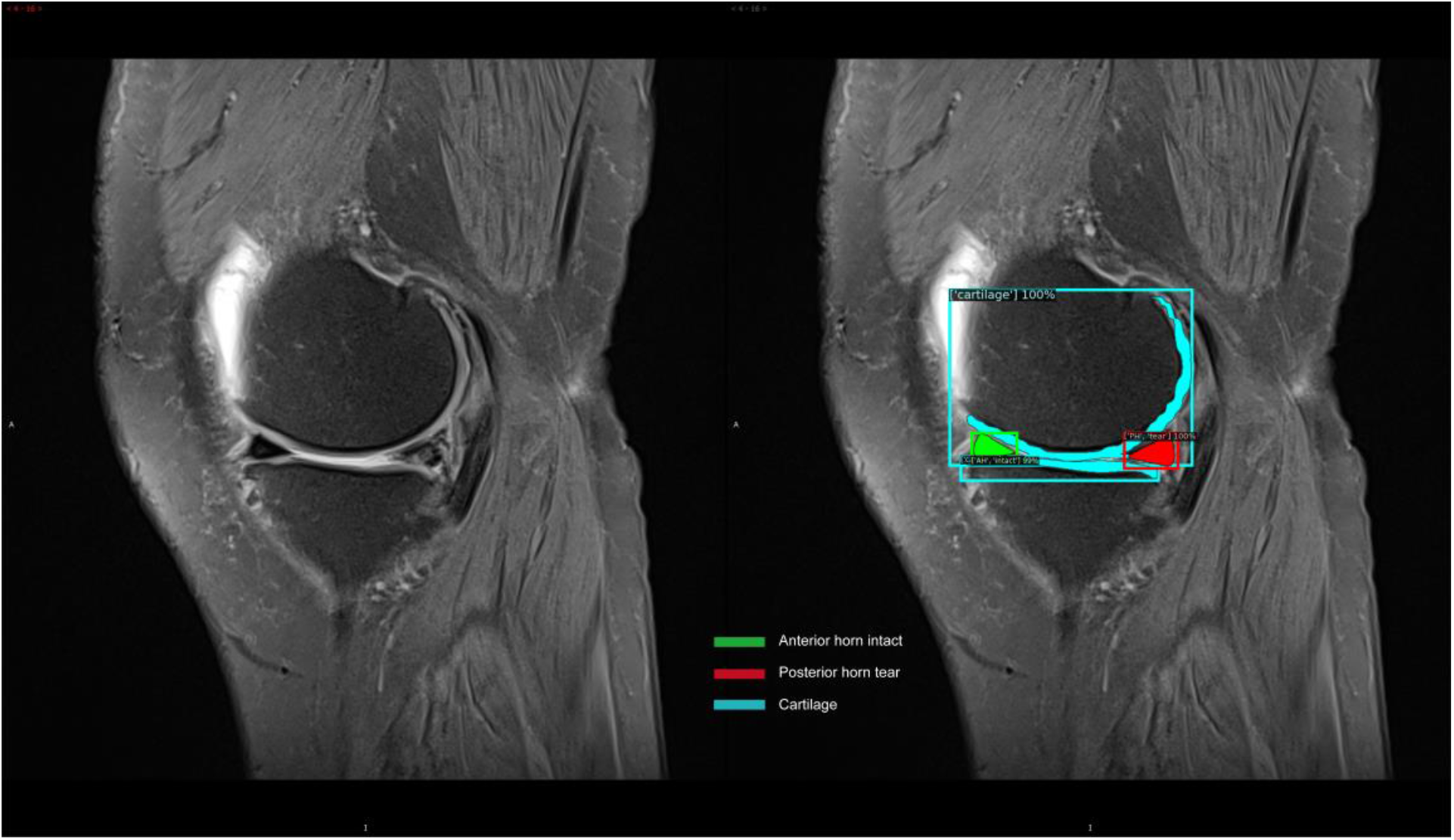
Classification and instance segmentation results of the meniscus MR images.

**Figure 6.**
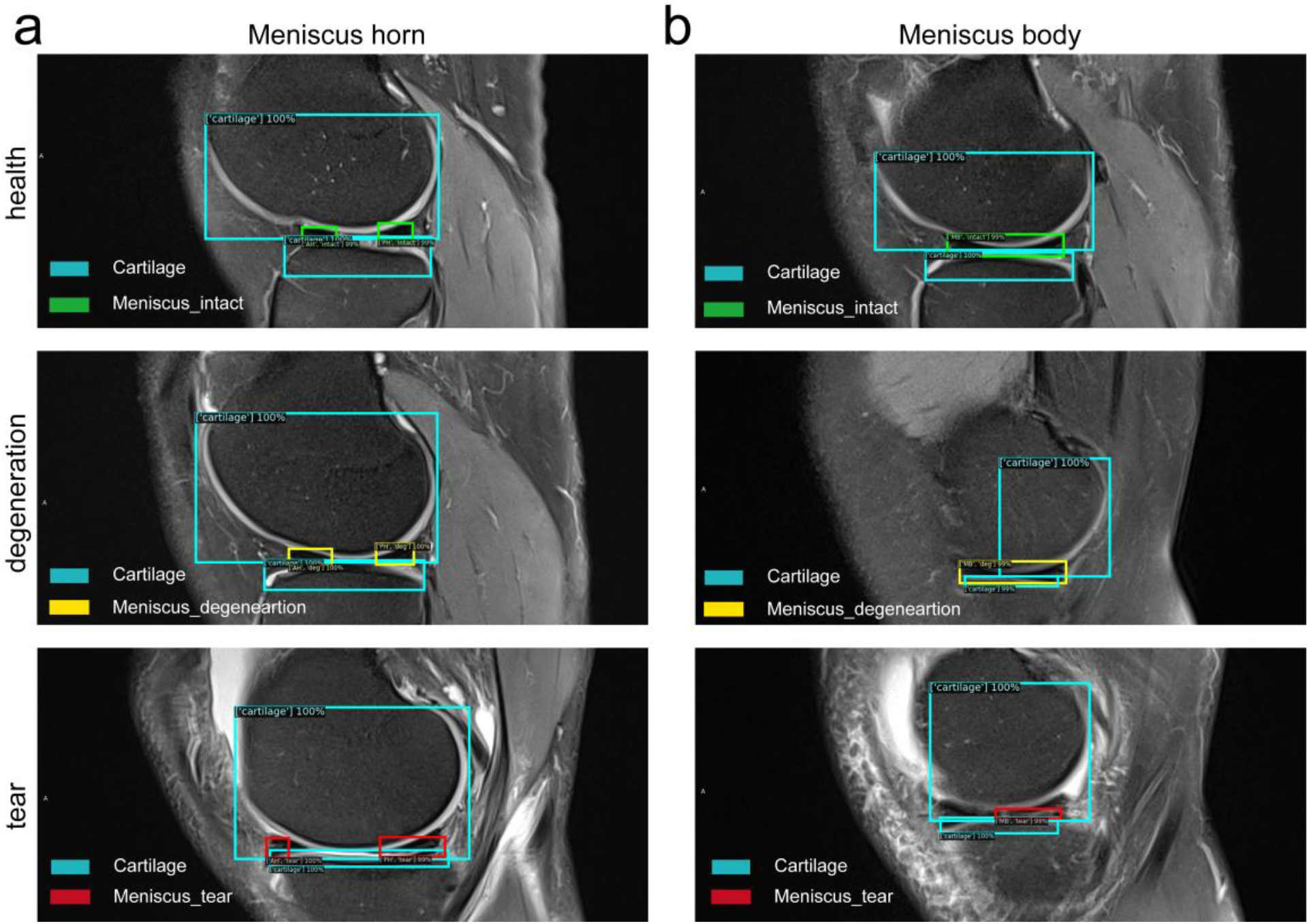
Bounding box diagnosis results of the meniscus MR images. (a) Meniscus horns, (b) Meniscus body.

**Figure 7.**
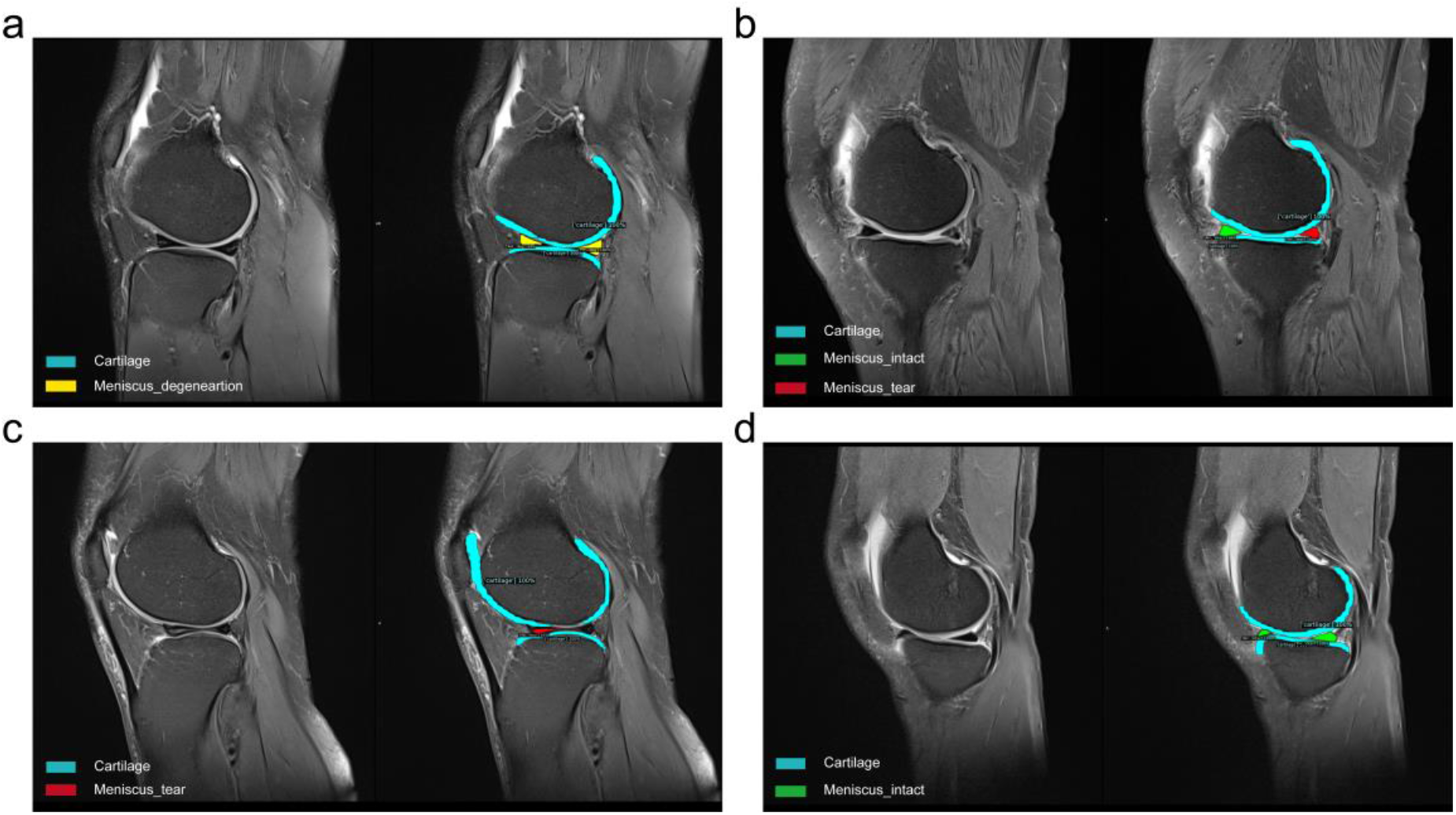
Mask diagnosis results of the meniscus MR images. (a) Degenerations at the meniscus anterior and posterior horns, (b) Tears at the meniscus body, (c) Tears at the posterior horn, (d) Healthy meniscus.

**Figure 8.**
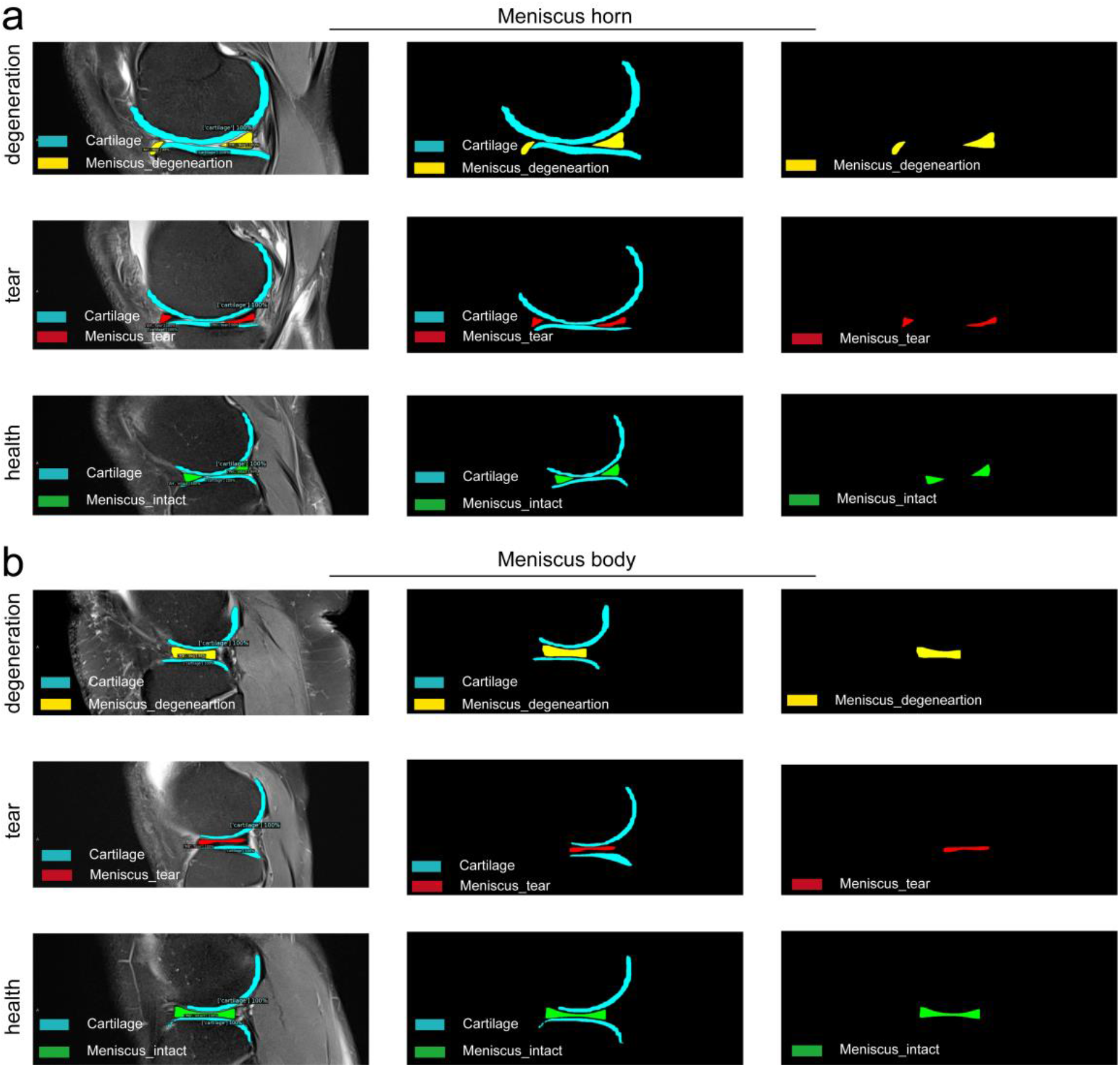
Diagnostic result highlighting and processing of the meniscus MR images. (a) Meniscus horns, (b) Meniscus body.

The Bbox diagnosis results demonstrated that the meniscus horns (**Figure 6a**) and the body (**Figure 6b**) were precisely divided into three categories: healthy, degenerated, and tear. The coverage of the cartilage tissue on each layer was also included in the Bbox. According to the Mask diagnosis results, the degeneration occurred at both anterior and posterior horns (**Figure 7a**), tears occurred at posterior horns (**Figure 7b**), tears occurred at the meniscus body (**Figure 7c**), and health meniscus (**Figure 7d**) was accurately identified.

The testing average precision of the Mask R-CNN with Resnet50_FPN as the backbone network is exhibited in **Table 3**. The results indicated that when the IoU shoulder was more than 0.5, the AP for the Bbox and Mask was 99.55 ± 0.41 %, 99.47 ± 0.28 %, respectively. As the IoU threshold value increased to more than 0.75, the AP for Bbox and Mask demonstrated a slight decrease. But the two values were more than 88%, which still exhibited extremely high accuracy. The AP for the different-sized objects were also acceptable, and all the values were more than 50%. Due to the deep network layers of the Resnet50, the identification APs were relatively good. For higher AP, the number of iterations was increased.

**Table 3.**
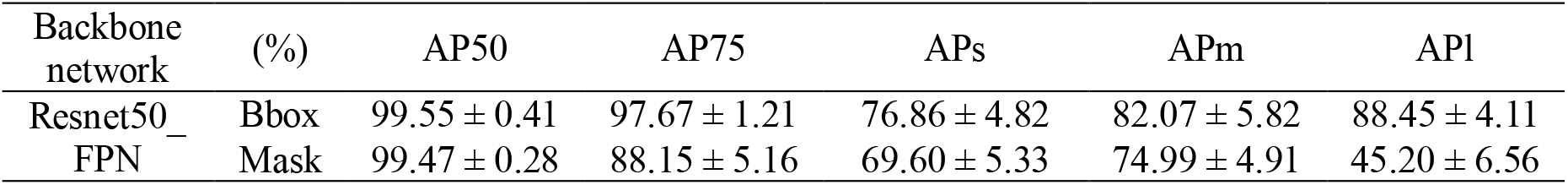
The AP of identification of the meniscus injuries.

### 3.3. The diagnosis of meniscus injures

**Table 4** represents the AP evaluation results of each category in the MRI images. The Bbox AP of the cartilage tissue was above 84%. The Bbox AP of the meniscus tear was more than 68%, and the value for the degenerated meniscus was more than 79%. As for healthy meniscus, the AP value was more than 80%. Although the mask diagnosis was a pixel-level diagnosis, the level of the AP values was similar to the Bbox AP values.

**Table 4.**
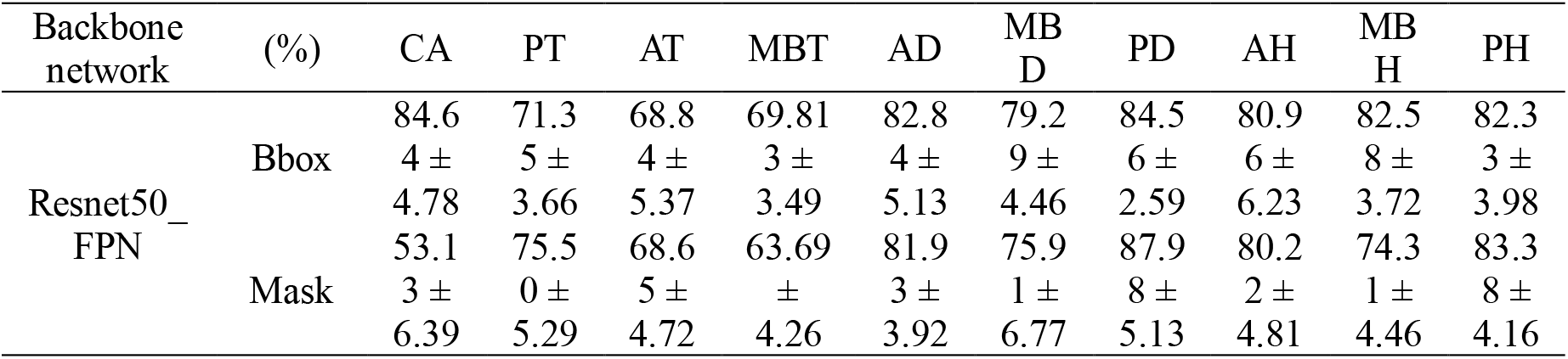
Per-category Bbox/Mask AP of identification of the meniscus injuries.

The sensitivity results at IoU from 0.50 to 0.95 are shown in **Table 5**. The overall sensitivity for the Bbox and Mask was 83.77 ± 5.29% and 74.43 ± 3.41%, respectively. The sensitivity for the target objects with different areas was also admirable. For the small and medium areas, the values were above 75%. For the large area, the Bbox sensitivity was as high as 95.77 ± 2.89%. Since the detection objects were relatively concentrated in the small and medium areas, the Mask sensitivity of large area scores was relatively low, but it still exceeded the critical value of 50%.

**Table 5.** Sensitivity of identification of the meniscus injuries.

| Backbone Network | (%) | Overall Sensitivity | Area =Small | Area =Medium | Area = Large |
| --- | --- | --- | --- | --- | --- |
| Resnet50_ | Bbox | 83.77 ± 5.29 | 78.16 ± 3.37 | 86.30 ± 5.28 | 95.77 ± 2.89 |
| FPN | Mask | 74.43 ± 3.41 | 73.54 ± 4.92 | 78.22 ± 4.36 | 59.67 ± 2.72 |

Compared to the diagnosis by experienced doctors, the identification and diagnosis accuracy were also quite high. Among the 200 patients of the internal testing dataset, 6 samples were unrecognized (3%). Besides, 49 of 56 healthy samples (unrecognized: 3 torn, 4 degenerated), 80 of 92 torn samples (unrecognized: 3 healthy, 9 degenerated), and 39 of 46 degenerated (unrecognized: 3 torn, 2 healthy) were recognized by the deep learning model. Therefore, the diagnosis accuracy for the healthy meniscus was 87.50%, for the torn meniscus was 86.96%, and for the degenerated meniscus was 84.78%.

For the external testing dataset, the 3.0T group demonstrated the better recognition rate and diagnosis accuracy than the 1.5T group (**Table 6**). Briefly, the healthy meniscus obtained the highest recognition rate in both 3.0T and 1.5T group. The lowest recognition rate occurred on the degenerated meniscus for both the two kinds of field strength. The torn meniscus can be effectively diagnosed in 3.0T group, but for the 1.5T group the diagnostic accuracy was the lowest among the three meniscus types.

**Table 6.** The verifying of the external dataset.

| Field strength | Meniscus type | Recognition rate | diagnostic accuracy |
| --- | --- | --- | --- |
| 3.0 | Intact | 93.33% (28 of 30) | 82.14% (23 of 28) |
|  | Degeneration | 76.67% (23 of 30) | 73.91% (17 of 23) |
|  | Tear | 86.67% (26 of 30) | 92.31% (24 of 26) |
| 1.5 | Intact | 80.00% (24 of 30) | 79.17% (19 of 24) |
|  | Degeneration | 66.67% (20 of 30) | 70.00% (14 of 20) |
|  | Tear | 76.67% (23 of 30) | 60.87% (14 of 23) |

The result of the arthroscopic surgery verifying was also optimistic, among the 40 diagnosed patients confirmed by the golden standard, 87.50% of them (35 of 40) obtained corrected diagnose using this model.

## 4. Discussion

The meniscus injury is one of the most common sports injuries. The MRI test generates a high soft-tissue image resolution, which is the first diagnostic choice for the meniscus injury [3]. But the diagnosis accuracy of the meniscus injury depends on the experience of the diagnostician. The popularization from the multi-center study on meniscus injury is hampered by the objective criteria of diagnosis, the subjective errors of doctors, and the diagnostic efficiency. These limitations put forward the objective demand for the standardization of the meniscus MRI images interpretation and the automation of classification. Herein, we proposed a deep learning network based on the Mask R-CNN to address the demands mentioned above. We used the most common conventional sequence and the routine scanning parameters, to ensure that it could be widely used in different hospitals, and the anatomical images from different MR machines were consistent. The results proved that this method could rapidly recognize the injured meniscus with high accuracy. Thus, we believed that this deep learning network can be regarded as an effective measure in the clinical application.

We used a deep learning method based on the Mask R-CNN to realize the meniscus injury identification and diagnosis. After the annotation and classification, meniscus MRI images of 924 individual patients were collected clinically. The substantial amount of images (nearly twenty thousands) were used in this study result in the admirable sensitivity and diagnostic accuracy. The meniscus images were segmented into 10 categories to estimate the meniscus injury. Compared to the similar studies involved the using deep learning model to diagnose the meniscus injury, this study has the largest label numbers (**Table 7**), indicated that the model can help the doctors to recognize more subjects on the MRI images. In the deep learning networks, the Resnet50_FPN was used as backbone networks, which had more network layers to integrate the features of the image at different depths. The RPN uses a 9-size anchor to extract the ROI from the original image, output category scores, and box scores, respectively. The Mask R-CNN finally delivered classification, bounding box regression, and mask diagnosis. While realizing the classification and recognition, the pixel-level diagnosis of the meniscus MRI injury was carried out. Through the training on the meniscus MRI image datasets, the AP of the bounding box regression was greater than 97%, and the AP of pixel diagnosis was greater than 88%. In the visualized diagnosis results, the size and location of the meniscus injury were displayed and marked, which was conducive to a better diagnostic estimate.

**Table 7.** The comparison of AI studies for meniscus tear diagnosis.

| Study | Reference standard | Label No. | Network structure | Sequences | Field strength | Image No. | Sensitivity |
| --- | --- | --- | --- | --- | --- | --- | --- |
| This study | Radiologists/<br>Arthroscopic surgery | 10<br>(intact/tear/degeneration/horn/body/cartilage//anterior/posterior) | ResNet | SAG FS PDW,<br>SAG FS T2 | 1.5/3.0 | 19872 | 83.77 ± 5.29% |
| Bien et al 11 | Radiologists | 2 (intact/tear) | MRNet | SAG T2, COR T1, ax PD | 1.5/3.0 | 1370 | 71% |
| Couteaux et al 29 | Radiologists | 4<br>(intact/tear/anterior/posterior) | ConvNet | FS-T2W | 3.0 | 1128 | AUC<br>Score=0.906 |
| Pedoia et al 31 | Radiologists | 2 (intact/tear) | U-Net | SAG 3D PDW | 3.0 | 1478 | 82% |
| Fritz et al 32 | Arthroscopic surgery | 2 (intact/tear) | DCNN | COR and SAG FS fluid-sensitive MRI | 1.5/3.0 | 20520 | 84% |
| Rizk et al 33 | Radiologists | 2 (intact/tear) | MRNet | SAG FS PDW, COR FS PD | 1.0/1.5/3.0 | 11353 | 89% |
| Roblot et al 30 | Radiologists | 3<br>(intact/horizontal tear/vertical tear) | Fast-RCNN/fast er-RCNN | SAG T2 | 1.5/3.0 | 1823 | AUC<br>Score=0.90 |

In the validating process of this study, the three types of menisci were well identified. The most common problem was the misidentification of a degenerative meniscus as a torn meniscus. According to the imaging principle, there may be two main reasons for this misidentification. The first reason is that the signal intensity, degeneration, and cleft meniscus are actually at the different stages of the same pathological process. The distinction between the two stages is not obvious, leading to the confusion of severe degeneration and spallation. The second reason is the complex anatomical morphology.

The meniscus is not regular in shape, its free edge is very slender with a thickness less than 0.5mm, and it is not easy to identify. In addition, when meniscal tears occur, broken fragments can shift to the femoral intercondylar notch and the original anatomic area of the meniscus is replaced by the fluid signals, which may result in erroneous recognition.

Except the validation on internal dataset, 180 samples from 8 hospitals scanned by MRI with different brands and field strength were used as the external dataset. The results demonstrated that this model have admirable recognition rate in 3.0T group, especially for the intact and torn meniscus. But the recognition and diagnose effect for 1.5T group was not as good as the 3.0T group. One possible reason is that the images obtained by the 1.5T MRI equipment were not employed in the learning and training process. The image quality including matrix and signal noise ratio differ exist between the 1.5T and 3.0T images. The recognition rate for degenerated meniscus was limited due to the ambiguous boundary between degeneration and tear. In addition, 40 samples who were diagnosed meniscus tear by the arthroscopic surgery were tested by this model to further verified the reliability, and the diagnostic accuracy rate was nearly 90%. Among the related studies, only one of them has employed the arthroscopic surgery as the reference standard. The introducing of this golden standard can improve the confidence of orthopaedic specialists in this model and promote the clinical application of this technique.

Artificial intelligence has a broad application prospect for the efficient analysis and classification of medical images. At present, there are great challenges in the application of artificial intelligence in diagnosing the abnormalities related to the knee joint. Except for the model algorithm and other technical factors, the biggest challenge is to establish a homogenized standard data set, and on this basis to adopt in different people and different performance MRI equipment. This requires a trade-off between diagnostic accuracy and universality. Herein this study, the training dataset was only collected from one 3.0T MRI equipment using the general scanning sequence. To enhance the learning and training effect, the data augmentation technique was used in the training process. This technique can simulate different slice angle, position, and fat suppression level by using geometric transformation, lighting adjustment, Gaussian filtering and noise addition. Through this operation, the dataset can be expanded by 20 times, meant that the images number has reached hundreds of thousands. But more importantly, the dataset can be further expanded by modifying the parameters of the currently used data augmentation methods.

In addition to improve the diagnostic accuracy, the AI technique also can be used to help doctors clearly distinguish the diagnosis. Herein, the image processing was performed to highlight the health of the meniscus. As shown in **Figure 7**, after removing the soft tissue background and cartilage, the deep learning model only recognized the situation of the meniscus. The green, yellow, and red pixels respectively represented healthy, degenerative, and torn meniscus. The horns (**Figure 7a**) and body (**Figure 7b**) of the meniscus, and the cartilage were easily recognized in this process.

There were several limitations in this study. First, only meniscus injury was identified, and no distinction was made between the types of meniscus tears. Moreover, the diagnostic accuracy of sagittal, coronal, and transverse views was not compared in the analysis. In the future study, we will analyze the accuracy of this model for the meniscus tears at different positions by including more cases and study the influence of the different layer thickness on the diagnosis of the meniscus tears using AI. Additionally, the validation method is also need to be improved. Only a few of the samples in this study was confirmed by the arthroscopic surgery, this result can partly reflect the effect of the deep learning model, but it is not statistically representative. In future research, it is necessary to strictly compare the arthroscopic diagnostic results with the data used for training and verification, so as to further improve the accuracy of the model.

## 5. Conclusion

In summary, a Mask R-CNN model was employed in this study to identify and predict meniscus tears on the MRI images. This deep learning model effectively detected and recognized the meniscus and cartilage, especially for tears that occurred at different parts of the meniscus. The recognition accuracy was more than 84%. With the increase of training sample size, the diagnostic accuracy can be further improved. The application of this technique can reduce the misdiagnostic rate of meniscus injury and alleviate the burden of doctors.

## Data Availability

All data produced in the present study are available upon reasonable request to the authors

## Abbreviations

MRI: magnetic resonance imaging
SE T1WI: spin-echo T1 weighted image
FSE T2WI: fast spin-echo T2 weighted image
FS FSE PDWI: fat-suppressed fast spin-echo proton density-weighted image
GRE: gradient echo
AI: artificial intelligence
PDW: proton density-weighted
CA: cartilage tissue
AT: anterior horn tear
PT: posterior horn tear
MBT: meniscus body tear
AD: anterior horn degeneration
PD: posterior horn degeneration
MBD: meniscus body degeneration
AH: anterior horn health
PH: posterior horn health
MBH: meniscus body health
RPN: region proposal network
ROI: region of interest
TP: true positive
FP: false positive
FN: false negative
AP: average precision
IoU: intersection over union

## Acknowledgement

This study was supported by the Natural Science Foundation of Jiangsu Province (BK20200121), China Postdoctoral Science Foundation (2020M671454), National Natural Science Foundation of China (81730067, 51575100), Nanjing Science and Technology Development Project (201803026).

